# Acceptability, Appropriateness and Feasibility of PROM and PREM Use in Swiss General Internal Medicine Divisions: A descriptive, sequential mixed method study

**DOI:** 10.64898/2026.04.29.26352033

**Authors:** Caroline Stan, Carole E. Aubert, Manuela Eicher, Jean Regina, Jérôme Stirnemann, Stefano Bassetti, Florence Vallelian, Lauren Clack, Vanessa Kraege, Marie Méan

**Author notes:** Author for correspondence: Caroline Stan, Faculty of Biology and Medicine, University of Lausanne, Rue du Bugnon 21, 1011 Lausanne, Switzerland.

## Abstract

**Background:** Patient-reported outcome measures (PROMs) and patient-reported experience measures (PREMs) are increasingly used to integrate patient perspectives into healthcare delivery, yet their routine implementation in general internal medicine (GIM) remains limited. This study evaluated participation rates and the acceptability, appropriateness, and feasibility of collecting PROMs and PREMs among GIM patients and study nurses across five Swiss university hospitals.

**Methods:** We conducted a sequential mixed-methods study embedded in a larger multicenter trial involving inpatients with two or more chronic conditions, hospitalized for acute illness and study nurses from GIM divisions. Inpatients completed three generic PROMs (paper or digital) at day 3, discharge, and 10 and 30 days post-discharge: the ESAS-r (Edmonton Symptom Assessment System revised), the EQ-5D-5L (European Quality of Life 5 Dimensions 5 Level), and the Distress Thermometer. A customized PREM assessing perceived quality of care was collected at discharge only. Patients and study nurses rated acceptability, appropriateness, and feasibility using Weiner’s implementation outcome measures. Study nurses’ recommendations for clinical integration were explored subsequentially in a focus group. Quantitative data were analyzed using descriptive analyses, while qualitative data were analyzed thematically.

**Results:** Among 1,773 eligible GIM inpatients, 59% (median age 72 years, IQR 63–81) agreed to participate in PROM and PREM collection. Overall, patients rated all the PROMs as highly acceptable, appropriate, and feasible. Study nurses rated the ESAS-r and the EQ-5D-5L accordingly but expressed a moderate rating for the Distress Thermometer and the PREM primarily for their ease of use. Focus group findings emphasized staff training, digital integration into electronic medical records, reduced questionnaire burden, and hierarchical support as key implementation facilitators.

**Conclusion:** Our study demonstrates that PROM and PREM collection in Swiss University Hospital Settings was considered acceptable, appropriate, and feasible by patients and study nurses in a multicentric GIM inpatient setting. Routine implementation warrants specific strategies.

**SUMMARY TABLE:** *What is already known on this topic:*

- PROMs and PREMs are widely used in many medical specialties to incorporate patient perspectives and evaluate the quality and value of care.
- Routine use of patient-reported measures in acutely ill GIM inpatients living with multimorbidity remains limited.
- Implementation often faces barriers related to workload, workflows, and digital infrastructure.

*What this study adds:*

- Two-thirds of acutely ill GIM inpatients in Swiss University Hospitals living with multimorbidity are willing to participate in PROM and PREM collection.

*How this study might affect research, practice or policy:*

- Staff training, digitalization, and hierarchical support are key facilitators, and embedding tools into electronic medical records with fewer measures may improve adoption in GIM.

## INTRODUCTION

The use of standardized patient-reported outcome measures (PROMs) and patient-reported experience measures (PREMs) has grown significantly in routine clinical care worldwide [1]. Implementation guidelines have been developed over the past decade [2,3], since PROMs and PREMs add a patient-centered dimension to healthcare evaluation. While PREMs serve as valuable performance indicators at different levels within health systems [4], use of PROMs, when integrated into structured patient follow-up, has been linked to better patient survival rates and reduced emergency department visits and hospitalizations [5]. Strategically, PROMs and PREMs enhance the comprehensiveness of hospital rankings [6] and are an effective means of addressing patient needs, thereby improving healthcare quality [7] and making them an important indicator for value-based healthcare.

However, the use of PROMs and PREMs remains fragmented and is often limited to specific fields. Previous studies reporting on the utilization of PROMs in medical patients mostly focused on specific populations, such as oncological [8–10], palliative [8], antiretroviral therapy-naive HIV infected [11], neurological [12], psychiatric [13,14], patients and on those with chronic disease [15–18]. One single study conducted among general internal medicine (GIM) inpatients in Norway showed a high participation rate and a significant symptom score variation between non-palliative and palliative patients [19].

The implementation process includes several steps, and numerous barriers to implementing PROMs and PREMs have been identified throughout. These include a) patient-related factors, such as the acute nature of illness, disease heterogeneity, multimorbidity, older age, and low digital literacy [20]; b) system-related factors, such as healthcare system fragmentation and limited digital integration during care transitions between in- and outpatient settings [21,22]. While these barriers are largely consistent across medical specialties, facilitators tend to be context-specific [23]. For instance, effective facilitators may include strong leadership support, interdisciplinary collaboration, dedicated training for healthcare professionals, and the availability of digital tools tailored to patient needs and clinical workflows [24]. Nurses have been identified as key stakeholders in PROMs and PREMs implementation, thus, patients and nurses perspectives are of particular interest [25].

We aimed to assess the acceptability, appropriateness, and feasibility rated by patients and study nurses of the use of generic PROMs and a customized PREM in patients hospitalized in GIM divisions across five Swiss university hospitals.

## METHODS

### Design & Setting

This sequential mixed-methods study assessed the acceptability, appropriateness, and feasibility of using PROMs and a PREM in patients hospitalized in GIM divisions. It included a quantitative assessment of participation rates and experience feedback among patients. For study nurses, it involved a sequential quantitative assessment of all three items, which informed the qualitative explanation and contextualization of the quantitative findings. The integration of quantitative and qualitative results occurred during the interpretation and reporting stages.

The study was part of a prospective, observational, multicentric cohort study performed in GIM divisions of five university hospitals (Bern, Basel, Geneva, Lausanne and Zurich; TRADUCE). It aimed to investigate the trends in the use of PROMs and a PREM among patients living with two or more chronic diseases hospitalized for acute illness in a GIM division. The TRADUCE study received local ethics committees’ approval from all five sites and is registered on the ClinicalTrials.gov platform of the National Institutes of Health (NCT06597318). Adult patients, capable of informed decision-making, of speaking French or German, and with a foreseen length of stay of over four days, were eligible to participate in the TRADUCE study that collected PROMs and a PREM.

Participating GIM divisions varied in size from 25 to 212 inpatient beds. The nurse-patient ratio is usually defined as 1:5-6, while working in multidisciplinary team approaches. None of the participating GIM divisions had experience of regularly collecting PROMs and PREMs.

The protocol for the present study received an ethics waver (CER-VD Req-2024-01078) as it falls outside the scope of the Human Research Act.

### PROMs and PREM

We used three validated measures to capture generic PROMs in French and German and one customized tool to collect a PREM. The Edmonton Symptom Assessment System-revised (ESAS-r) [26–28] assesses ten symptoms commonly experienced by patients with advanced disease, using a 0 to 10 scale. The EQ-5D-5L index [29] measures health-related quality of life in five dimensions. The Distress Thermometer (DT) [30] quantifies psychological distress, with defined cut-offs. The PREM consisted of a short questionnaire with open- and closed-ended questions that allowed patients to comment on their experience of the quality and quantity of care received at the end of their hospital stay (**Supplementary Material 1**). The questionnaire was based on previous versions [31] and Swiss Association for Quality (ANQ) quality-of-care items [32]. Patients were followed according to a structured data collection protocol, with admission and follow-up visits performed at five key time points: Day 0, Day 3, hospital discharge, Day 10 post-discharge, and Day 30 post-discharge (**Supplementary Material 2)**. Three hospitals gave patients a choice of format, one offered only digital, and one offered only paper. For paper measures, study nurses gave patients the measures at their bedside for inpatient collection and mailed them to them for outpatient collection. The process was carried out remotely for digital measures.

### Data collection

Data collection took place between September 2024 and February 2025. We re-used routinely collected demographic and clinical data (age, sex, comorbidities, medication during hospital stay, rehospitalization and emergency division visits). The PROMs and PREM data were recorded in local systems specific to each hospital, using a combination of paper and digital media, which were then integrated into each hospital’s local data warehouse (DWH) [33]. Each hospital used a different data collection system; details are provided in **Supplementary Material 3.**

### Data analysis

#### Patient perspectives

We first assessed how many patients agreed to participate in the study and to use PROMs and a PREM. The patient participation rate was determined by comparing the number of patients who agreed to participate among those screened and eligible for the study. Reasons for declining participation in the use of PROMs and PREM were categorized into three main themes: (1) lack of interest; (2) overall burden; and (3) burden related to follow-up. These reasons were documented by research collaborators at each participating hospital. The frequency of digital versus paper formats, as well as follow-up completion rates, were compared across sites.

To gain patient perspectives, patients were contacted by phone 30 days after hospital discharge and invited to share their opinions on the use of PROMs and PREM. One hundred consecutive patients—representing 10% of the total sample—participated in this part. The telephone interviews, lasting approximately five minutes, were conducted using a validated set of implementation outcome measures: the Acceptability of Intervention Measure (AIM), the Intervention Appropriateness Measure (IAM) and the Feasibility of Intervention Measure (FIM) [34]. These scales were designed and validated to measure the extent to which participants perceive interventions as acceptable, appropriate, and feasible. As described in this previous publication, acceptable refers to the perception that an intervention is agreeable, appropriate to the perceived fit of an intervention in a given setting, and feasible to how an intervention can be successfully used in a setting [34].

This set of instruments was designed to assess three key aspects of an intervention – in this case the overall use of PROMs and PREM of the TRADUCE study. Each of the AIM, IAM, and FIM consists of four statements that are rated on a 5-point Likert scale, assessing acceptability, appropriateness and feasibility of an intervention, respectively. It was available in English and German, but not in French (**Supplementary Material 4**). To ensure its relevance and comprehension among French-speaking patients, the validated English one was translated into French, following ISPOR guidelines (**Supplementary Material 5**) [35]. Descriptive quantitative analyses of the responses were performed.

### Study nurse perspectives

Five study nurses, one per participating hospital, who were directly involved in patient recruitment, data entry, and follow-up processes related to the use of the PROMs and PREM, were invited to complete an online survey (**Supplementary Material 6**), which included twelve Likert-scale questions designed to assess agreement and satisfaction levels. The survey covered feasibility, barriers and facilitators, usefulness and impact, digitalization, follow-up, study execution, and professional background. Study nurses completed the AIM, IAM, and FIM [34] separately for each measure (ESAS-r, EQ-5D-5L, DT and the customized PREM).

Data from the online survey responses were used to guide the design and inform the semi-structured focus group in English. The purpose of the focus group was to explore in greater depth the key themes identified in the questionnaires, thereby generating richer qualitative insights. Discussion topics included the advantages and disadvantages of the intervention, implications for clinical practice, and barriers and facilitators to implementation. The focus group was facilitated by the first author following a structured guide (**Supplementary Material 7**). Lasting approximately one hour, the session was recorded and subsequently analyzed.

Data from the focus group were analyzed thematically using the Braun & Clarke method [36], to identify key patterns and insights related to the use of the PROMs and PREM. The data analysis followed six steps: familiarization with the data, generating initial codes, searching for themes, reviewing themes, defining and naming themes, and finally producing the report. The coding process was primarily inductive using MAXQDA® 2024 (VERBI Software, 2024). The analysis and presentation were then structured around overarching themes derived from prior survey responses, used to guide the focus group and produce a predefined analysis grid (**Supplementary Material 8**), with titles corresponding to the themes.

## RESULTS

### Patient perspectives on the use of the PROMs and PREM

#### Participation rate

Overall, 1,773 patients were screened and 1,052 (59%) accepted to participate. Overall, participating patients were 41% (436) female with a median age of 72 years [interquartile range (IQR): 63 to 81].

Of the 721 patients who declined to participate, 59% (423/721) cited lack of interest, 38% (275/721) mentioned the overall burden of filling in measures, and 3% (23/721) reported follow-up burden. Participation rate in hospitals ranged from 47 to 75%. The highest participation rate was observed at Hospital 1 (**Supplementary Material 9**).

#### Digital vs paper format for the PROMs and PREM

A total of 470 participants (45%) used the digital format. By excluding the two hospitals that offered only one option (paper or digital only), there were a total 511 participants, with 179 (35%) responding digitally and 332 (65%) on paper. The proportion of digital use varied substantially among the hospitals that offered a choice of format, ranging from 9% to 76%. Median age was 64 years (IQR: 55-74) for the digital format versus 73 years (IQR: 65-82) for the paper format (**Figure 1**).

**Figure 1.**
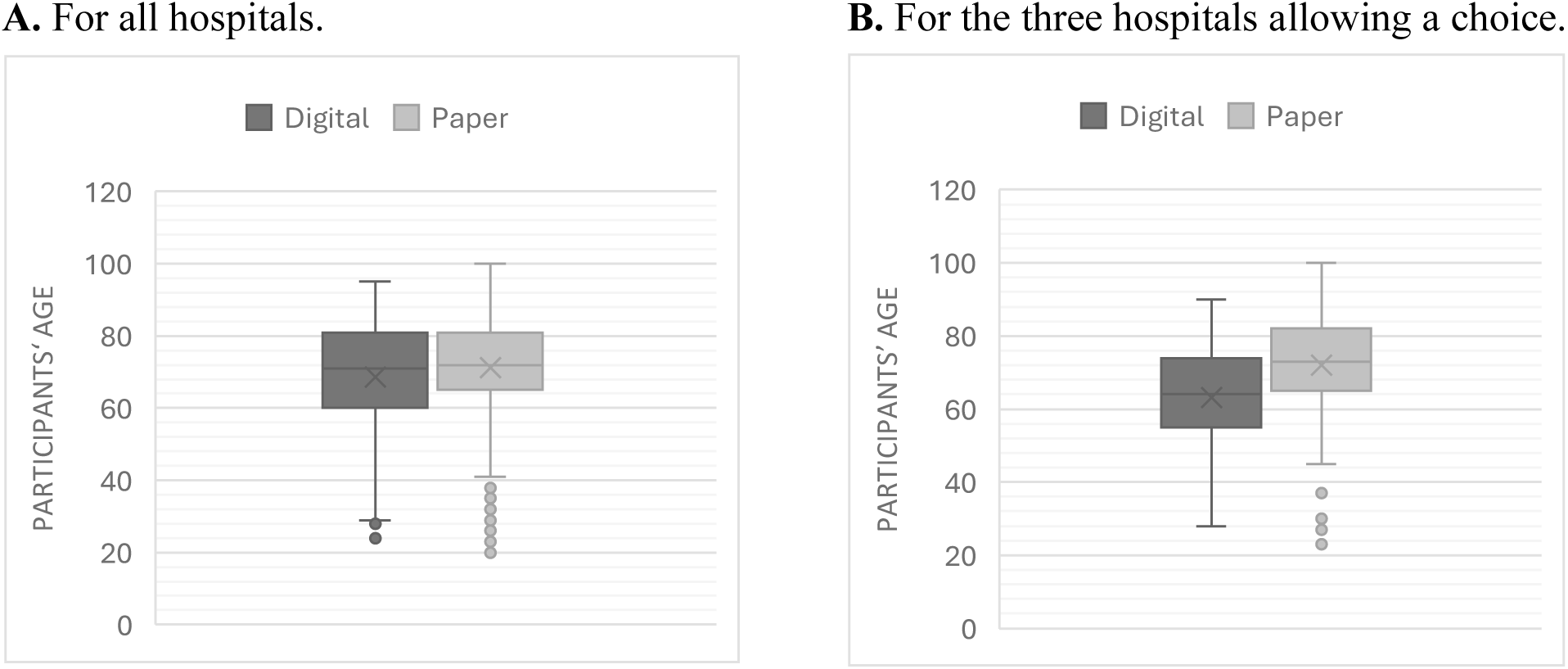
Age distribution by format of measure use.

#### Acceptability, appropriateness, and feasibility of PROMs and PREM

Overall, more than 70% of patients (combining “agree” and “completely agree”) rated the reporting of PROMs and PREM as acceptable, more than 74% as appropriate, and more than 78% as feasible (**Table 1**).

**Table 1.**
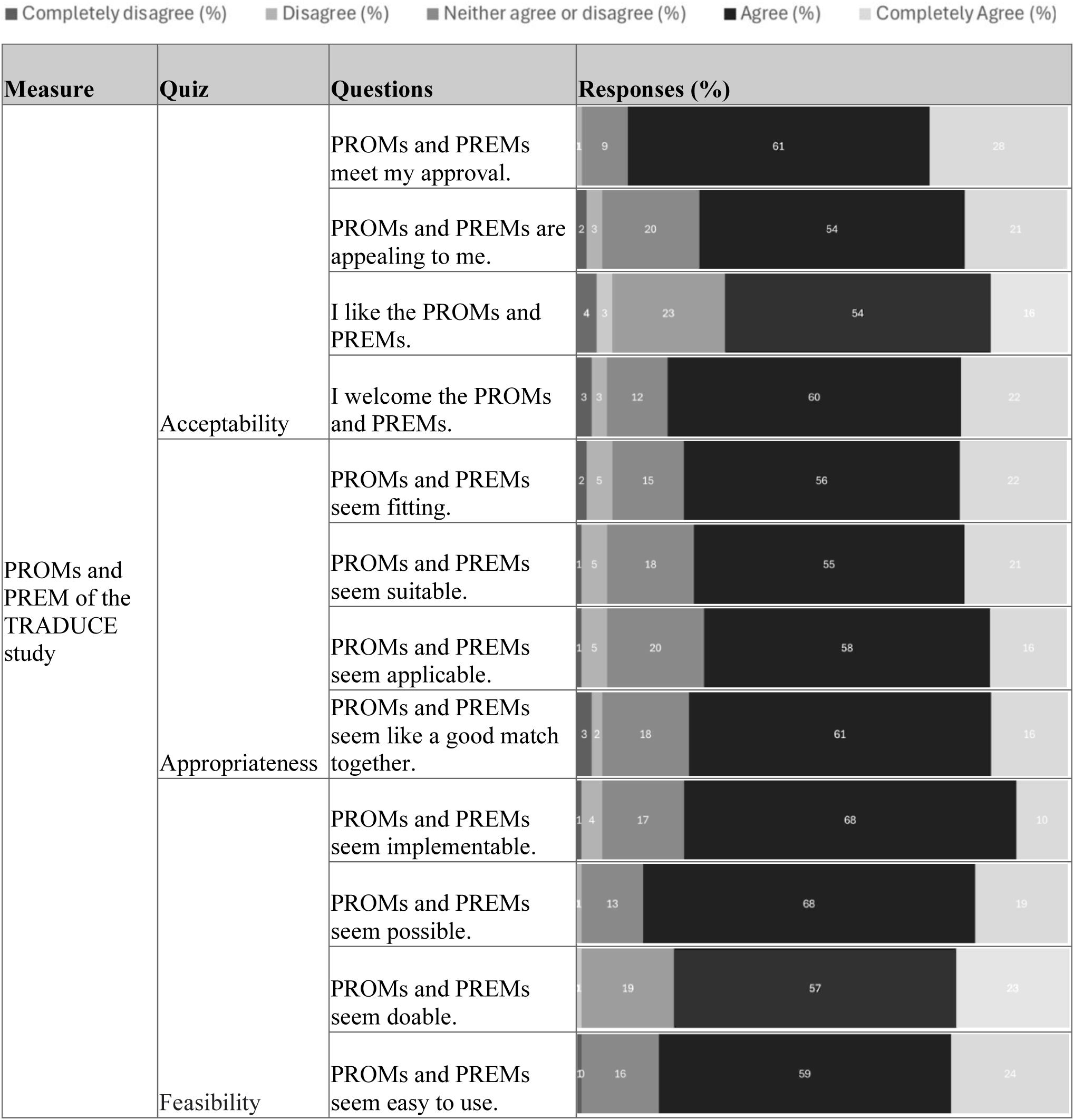
Responses of the patients to the acceptability, appropriateness and feasibility of intervention measures.

### Study nurse perspective on the use of the PROMs and PREM

#### Participation rate

All five study nurses, one from each participating hospital, responded to the AIM, IAM and FIM and participated in the online survey and semi-structured qualitative focus group.

#### Acceptability, appropriateness, and feasibility of PROMs and PREM

Most study nurses rated the PROMs (ESAS-r, EQ-5D-5L, and DT) and the PREM as acceptable, appropriate, and feasible (**Table 2**). High levels of agreement were observed for the ESAS-r and EQ-5D-5L, although one nurse disagreed with the ESAS-r’s ease of use. Responses for the DT were more variable, with acceptability and appropriateness ranging from neutral to positive, and feasibility, particularly ease of use, receiving mixed ratings. The PREM was generally viewed positively, though some negative ratings were reported, primarily concerning acceptability and ease of use (**Table 2**).

**Table 2.**
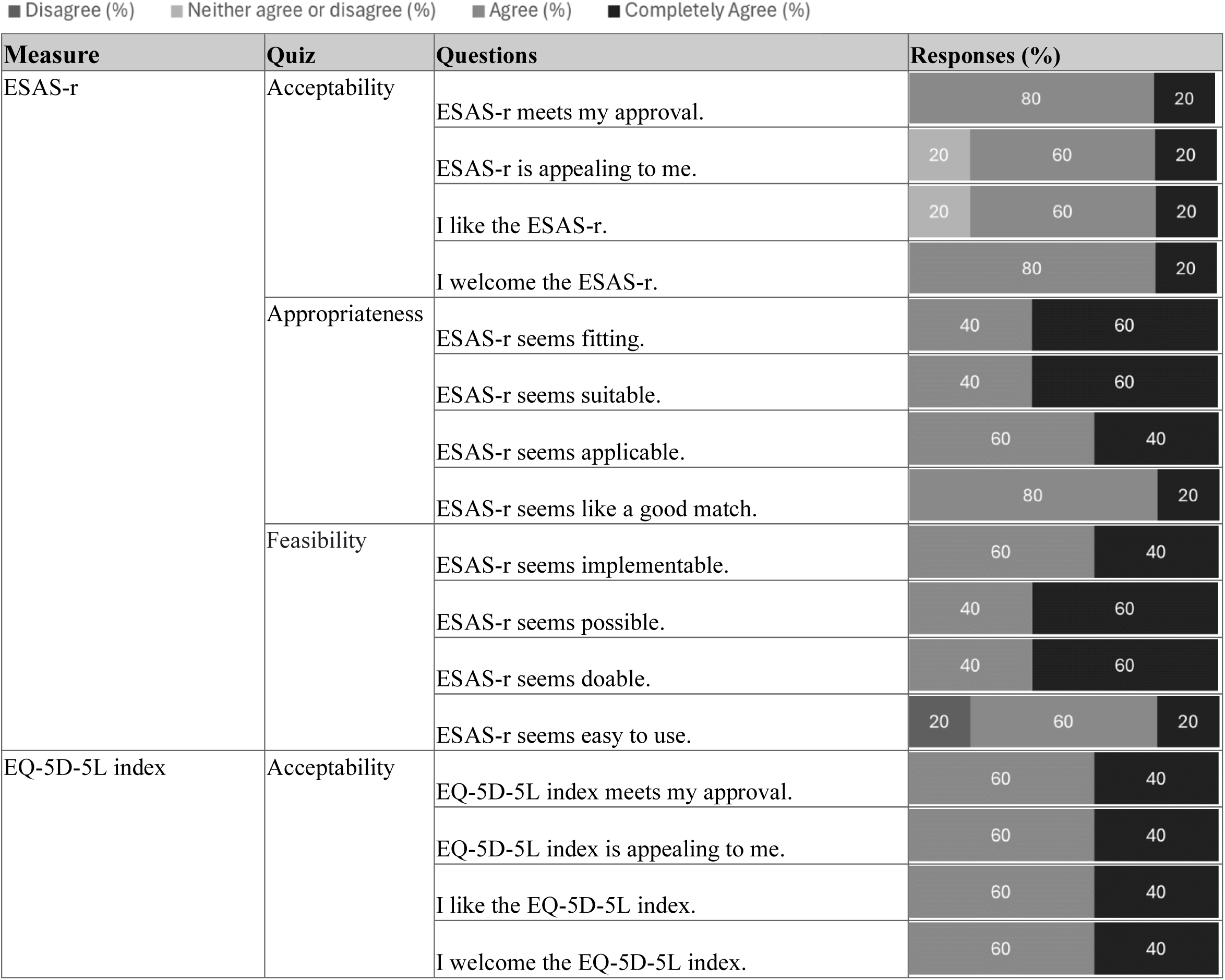
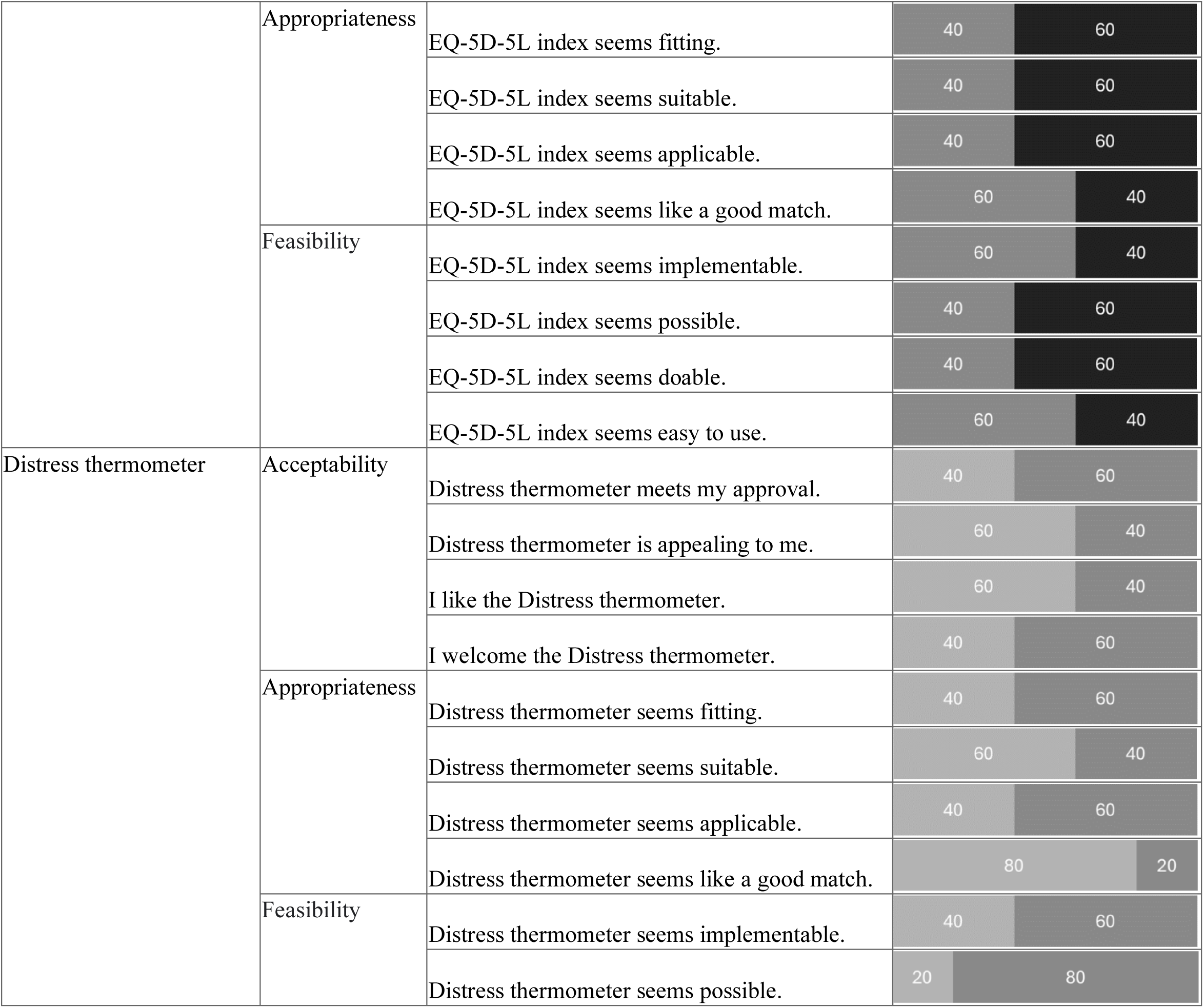
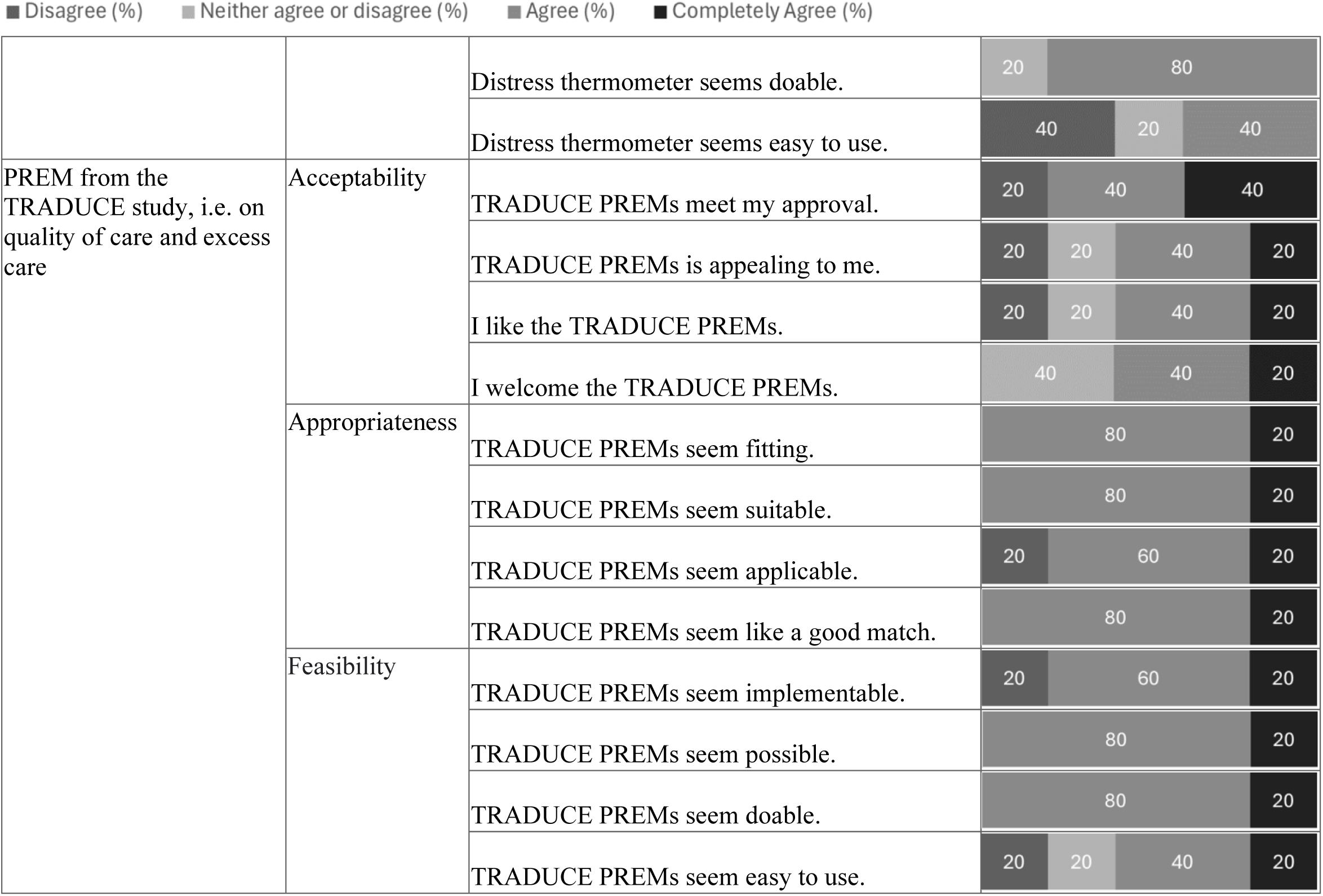
Responses of the study nurses to the online survey – acceptability, appropriateness and feasibility of intervention measures.

#### Questions of the online survey

Study nurses emphasized that PROMs and PREM were most suitable for certain specific pathologies but offered less flexibility for GIM patients living with multimorbidity. A major barrier was the low user-friendliness of digital tools and the burden and time required to access them, keeping paper measures common, especially for older patients. Challenges included incomplete or incorrectly filled measures and declining patient motivation after discharge. Additional barriers identified in the survey were lack of resources (4/5), resistance to change (2/5), and a fragmented care system (1/5) (**Table 3**).

**Table 3.**
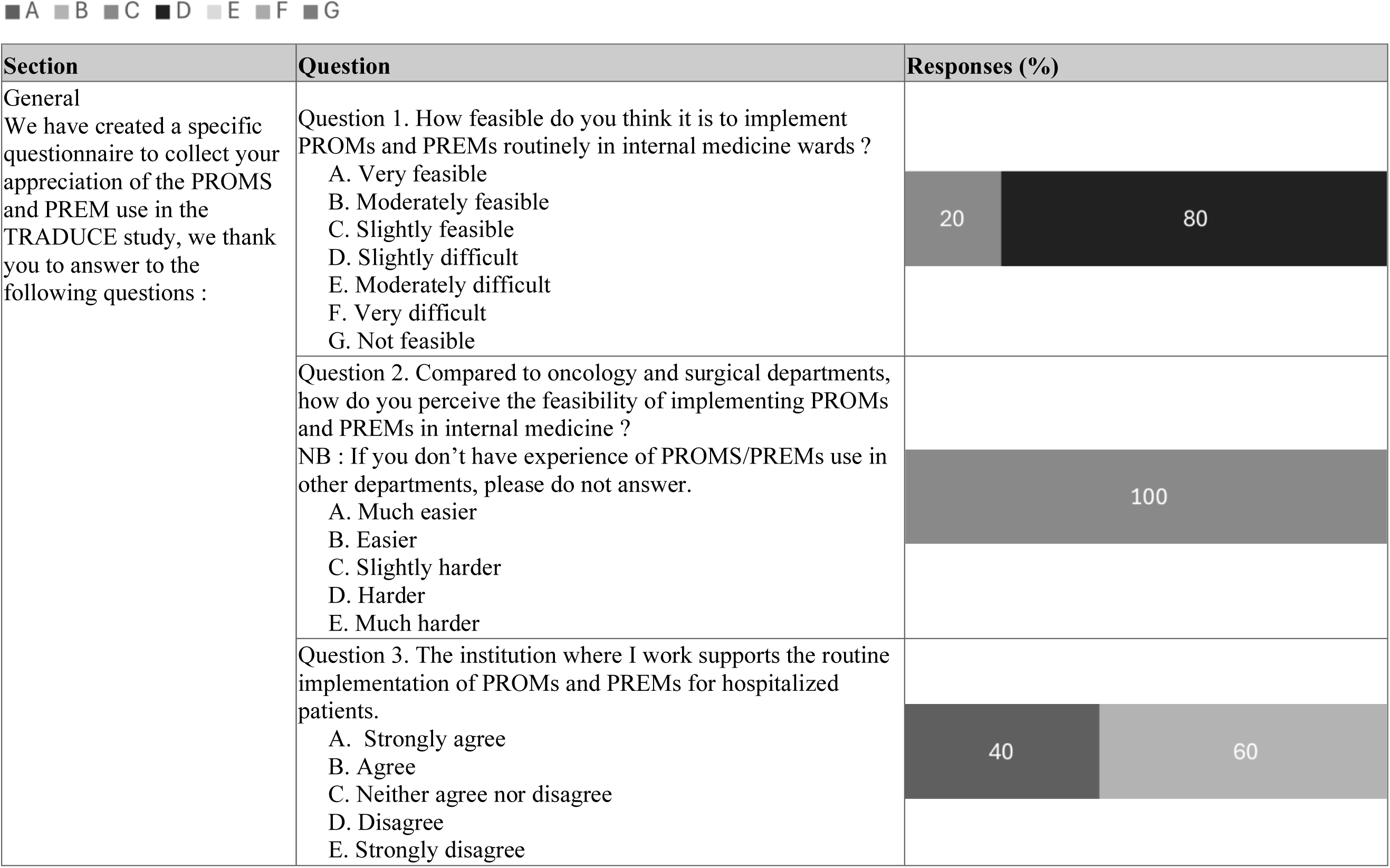
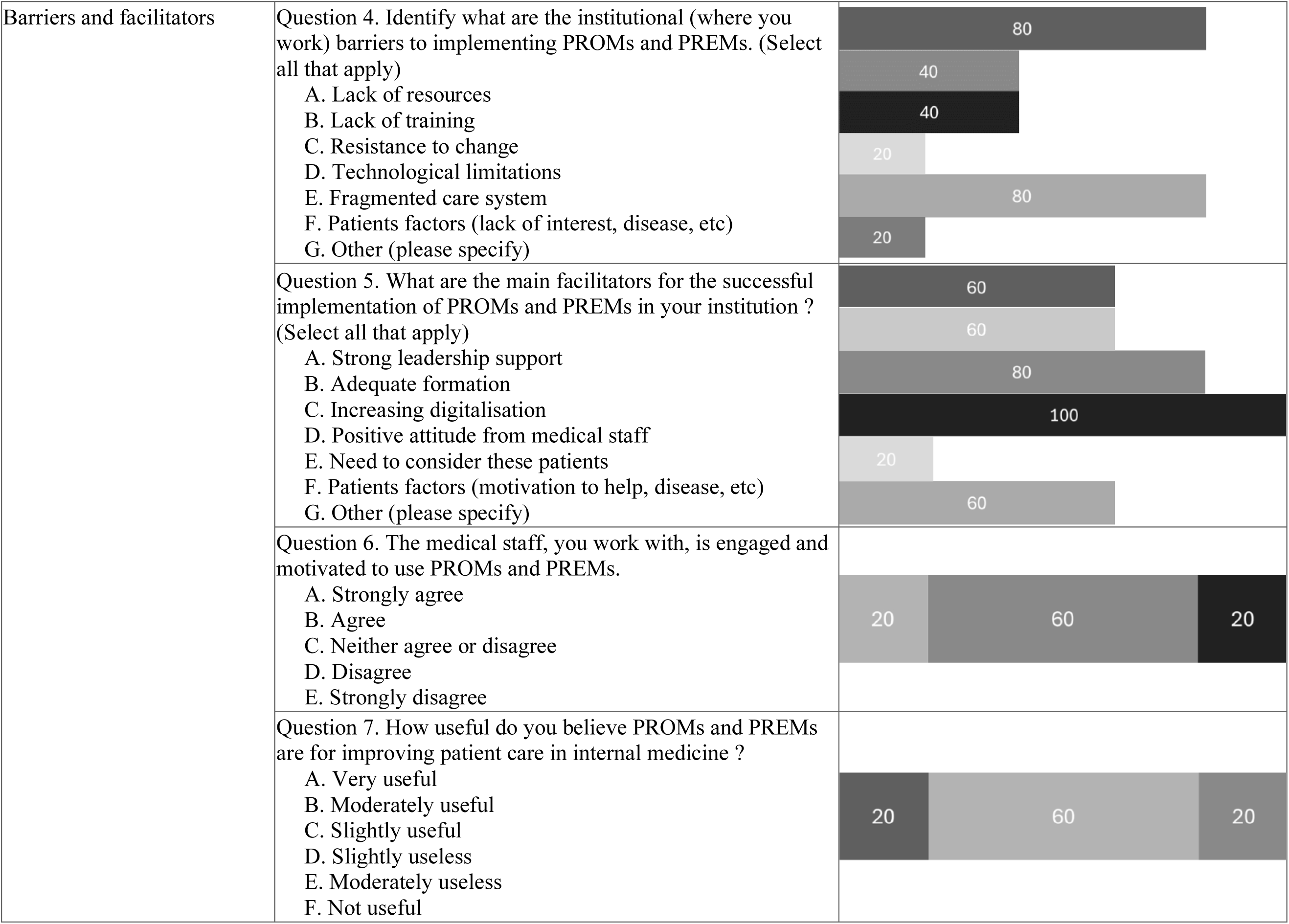
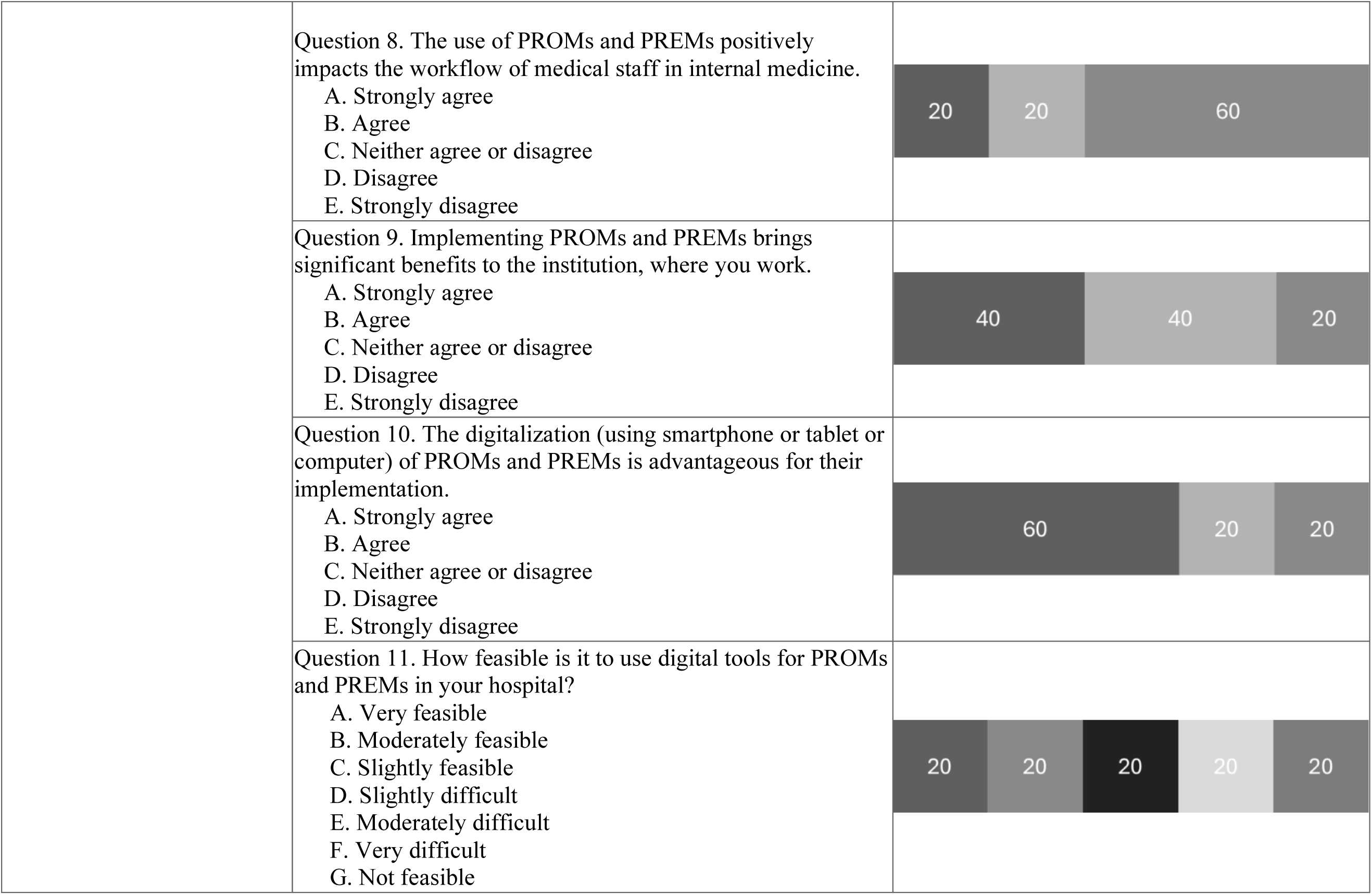
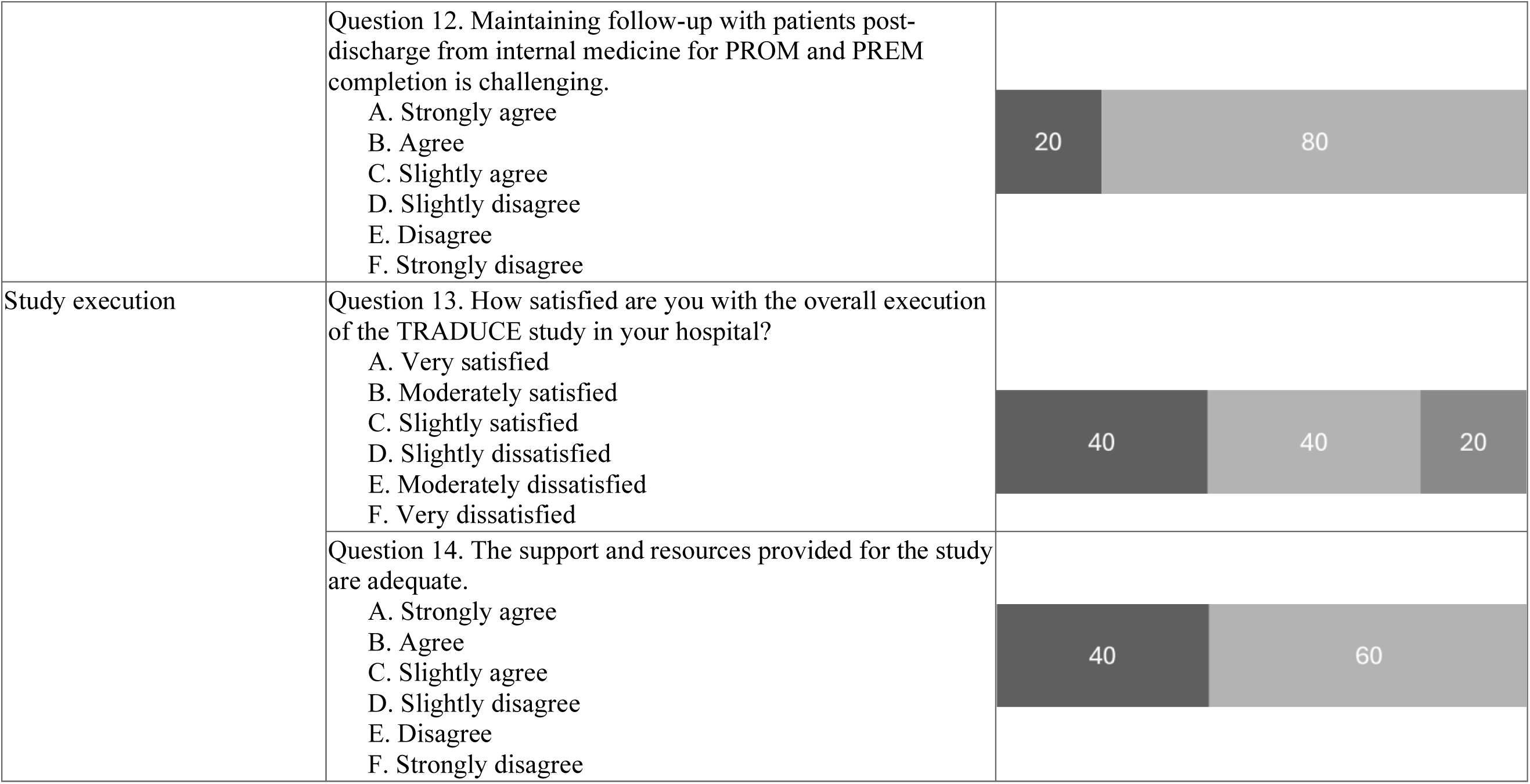
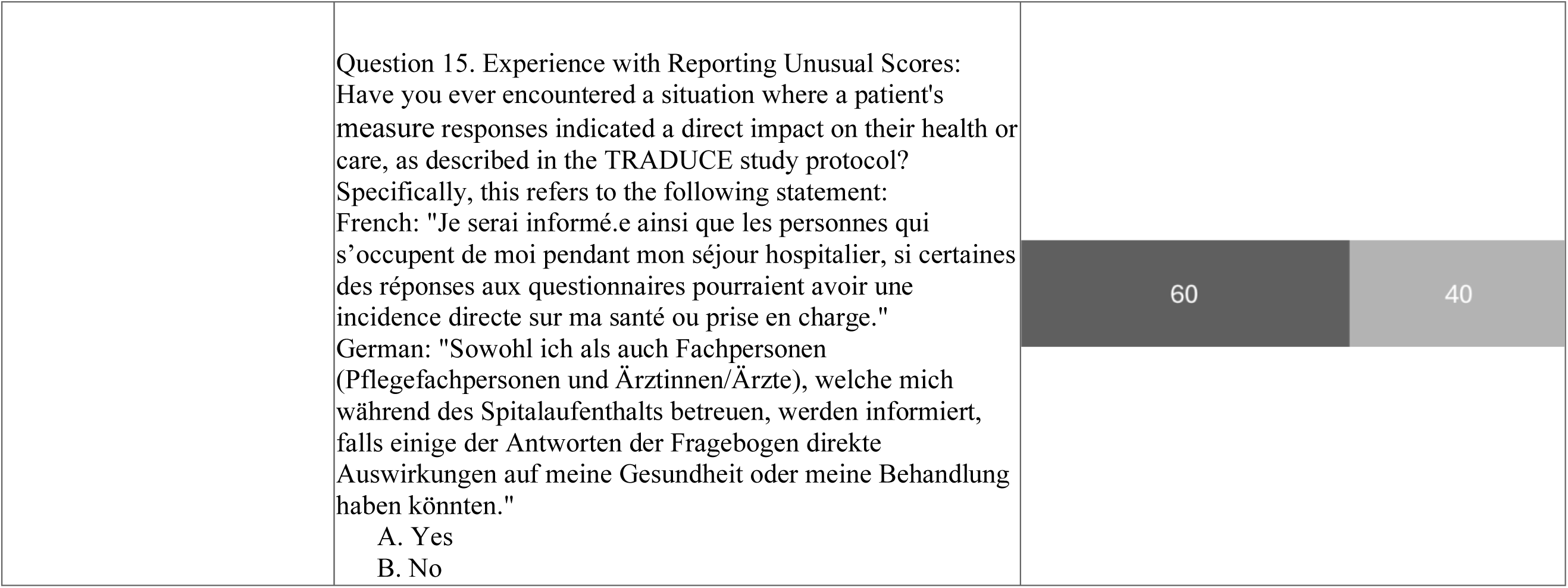
Responses of the study nurses to the online survey – closed-ended questions.

Suggestions for improvement included reducing the number of measures, especially at discharge, and clarifying the wording of items to avoid confusion. The use of shorter and better adapted versions of the tools, such as the EQ-5D-5L, was mentioned. Facilitators included positive staff attitudes (5/5), increasing digitalization (4/5), management support (3/5), adequate training (3/5), patient engagement (3/5), and recognition of patient perspectives (1/5).

#### Semi-structured qualitative focus group

The data reflects study nurses’ perspectives, including their subjective impressions of patient experiences. A detailed presentation of the qualitative findings is provided in **Supplemental Material 10**. Schematic summaries of the qualitative analysis are illustrated in **Figure 2**.

**Figure 2.**
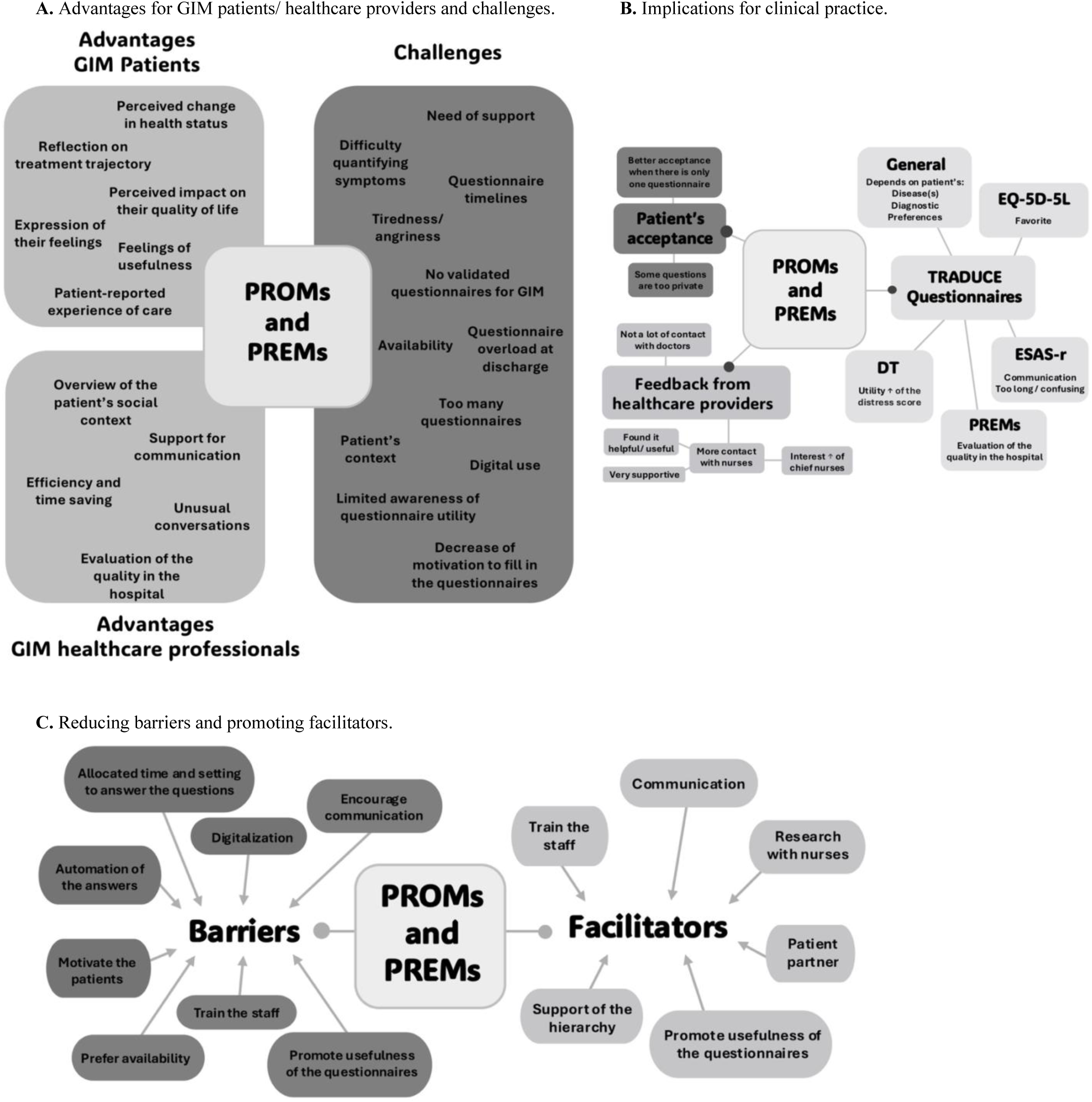
Results of the thematic analysis of the focus group with the study nurses **Foot note:** PROMs (Patient-Reported Outcome Measures); PREMs (Patient-Reported Experience Measures); GIM (General Internal Medicine); ESAS-r (Edmonton Symptom Assessment System-revised), EQ-5D-5L (European Quality of Life 5 Dimensions 5 Level), DT (Distress Thermometer)

##### 1. The impact of PROMs and PREMs for patients

Study nurses unanimously recognized the value of these measures for patients. They noted that PROMs and PREMs help patients better understand their health, track the evolution of symptoms, and feel more engaged in their care by providing an opportunity to express their feelings. These tools were described as facilitating communication, enabling discussions on issues that might not arise during conventional consultations, and potentially saving time by making appointments more efficient.

However, several challenges were identified. Some patients, particularly older people or those with multiple comorbidities, struggle to complete the measures, especially digital versions. Certain questions, especially those addressing sensitive topics such as mental or sexual health, were considered intrusive by some patients.

The ESAS was described as useful for communication, although lengthy and occasionally confusing. The EQ-5D-RL was considered particularly relevant for assessing patients’ current quality of life (study nurse 3): *« It’s really [about] how they live with their disease and how it impacts their health. » (minor editorial modifications made for clarity).* The DT, on the other hand, received mixed reviews: it was cited as useful for identifying psychological distress in some patients, but some found it intrusive, especially regarding mental or spiritual health (study nurse 2): *“I think sometimes the questions were very private, especially during moments of distress.”* Regarding PREMs, study nurses highlighted their role in evaluating patients’ perception of care quality (study nurse 1): *“I think PREMs are also useful for quality management, to demonstrate whether the quality of care in the hospital is adequate.”*

##### 2. Barriers and facilitators to implementation in GIM divisions

This theme highlights the main barriers and facilitators to implementing the PROMs and PREM in GIM divisions. Key barriers included difficulties faced by older patients in using digital tools, often requiring support to complete measures and struggling with different rating scales. Study nurses also noted a loss of patient motivation, particularly after hospital discharge, and challenges in finding convenient times for completion (study nurse 3): *“With the digital questionnaire, it’s easier. With the paper one, I encountered issues with patient availability—they are with nurses, physiotherapists, interns, or attending to personal needs such as toileting or showering.”* Digitization was highlighted primarily as a facilitator.

##### 3. Study nurse suggestions for the implementation of the PROMs and PREM

This theme explores professionals’ insights into integrating PROMs and PREMs into routine care. Proposed solutions include training healthcare staff, improving communication about their benefits, and integrating them into electronic medical records to reduce burden and simplify data collection (study nurse 4): “*It should be recorded in one place, maybe with a score or something else, that the healthcare team could see*.”

Other suggestions included reducing the number of measures and better scheduling of patient appointments to ensure they are available to respond accurately, i.e. without distractions or noise (study nurse 1): “*Patients can reflect more on their answers because they do it alone*.” Training and awareness for both staff and patients were emphasized as essential.

## DISCUSSION

This sequential mixed-methods study, collecting feedback on PROMs and a PREM from 1,052 patients living with multimorbidity hospitalized for acute illness and from five study nurses across five Swiss GIM divisions, demonstrates that these tools are generally acceptable, appropriate, and feasible from the perspectives of both patients and study nurses. Two-thirds of patients were willing to participate, and nearly half completed digital versions of the measures. Patient responses showed high agreement across acceptable, appropriate, and feasible items, while study nurses’ assessments were more tempered, particularly regarding the DT. Nonetheless, overall feedback was predominantly positive. In the focus group, study nurses highlighted staff training, digitalization, and hierarchical support as critical for integration into routine practice. Key recommendations to overcome implementation barriers included embedding measures in electronic medical records, reducing their number to lower patient burden, and enhancing communication about their benefits.

The participation rate of 59% is in accordance with the 64% reported by Neve et al. in an otolaryngology practice in the Netherlands [37], and the 67% in Swiss cardiac rehabilitation programs, a population often similar in terms of age and comorbidity to the population of our study [38].

The most frequently reported reason for non-participation was a lack of interest. Similarly, during follow-up after discharge, many patients were unable to participate due to poor health status. A prior study conducted in a comparable GIM setting in Norway reported similar findings, noting that patients in poorer clinical condition and therefore unfit to participate represented a significant challenge [19]. This challenge has also been reported in postoperative follow-up, where only a minority of patients are willing or able to continue completing PROMs one year after surgery [39].

In the present study, 35% of patients given the choice, agreed or expressed willingness to use the digital format. These findings align with a study conducted in the Netherlands in a colorectal cancer setting, in which only 41% of respondents in the paper-optional group opted for electronic data collection [40]. In Switzerland, digital skills of the population are above the OECD international average in eHealth literacy, numeracy and adaptative problem solving [41]. However, according to the MONET 2030, a Swiss indicator designed to assess the current situation and development in Switzerland, only a limited share of people has advanced digital competencies, or skills beyond basic use [42]. There are especially large gaps among older adults. Our study confirmed an age-related preference and accessibility pattern, with patients favoring paper-based questionnaires. This was reflected in the median ages of users by format: 64 years for digital and 73 years for paper.

Patients demonstrated a high level of agreement regarding the acceptability, appropriateness, and feasibility of the PROMs and PREM used in the present study. In particular, the tools were rated highly for their ease of use and perceived practicality. These findings are consistent with the PRO Heart-DK study, in which over of 75% of cardiac rehabilitation patients reported PROMs to be highly acceptable and useful for preparing clinical consultations and enhancing communication [43].

The **ESAS-r** received positive overall ratings from the study nurses. While being criticized for being lengthy and confusing at times, its usefulness for communication was underlined in the focus group. A review conducted within the Ontario palliative care context emphasizes its effectiveness in improving symptom detection, control, and patient satisfaction [44]. Ontario palliative care experts recommend team education and training to improve their use.

The **EQ-5D-RL** was the best ranked among all dimensions and was considered particularly relevant for assessing patients’ quality of life by study nurses. In the context of patients with chronic diseases, its speed and ease of use were emphasized [45]. Regarding its potential of use, a longitudinal study of people undergoing palliative home care has shown that it could be used as an initial tool to quickly identify needs and guide clinical decisions. However, its use in GIM may be better suited to quality improvement initiatives or research, given its generic nature, that limits its ability to detect changes specific to a condition or symptom [46].

The **DT** was considered challenging to use by study nurses. This correlates well with existing literature. A study conducted in Switzerland reported logistical issues with administering the DT, mostly occurring on the day of admission when patients and nurses tend to be busy. The nurses in the study reported other barriers, including high staff workloads and a lack of reminders in the electronic health record [47]. However, the mentioned barriers do not seem to be population specific. Despite these practical limitations, evidence demonstrates that the DT enables the identification of patients with social needs in an acute inpatient oncology setting, facilitates timely referrals to appropriate support services, and contributes to more appropriate utilization of healthcare resources [47].

### Clinical implications

Regarding the future implementation of the PROMs and PREM in GIM divisions, digitalization was perceived by study nurses as a moderate barrier. Limited digital literacy may hinder both acceptance and effective use of digital tools in clinical practice. Nevertheless, digital formats were also viewed as offering important advantages, including more precise responses, reduced missing data, and greater flexibility, by allowing patients to complete measures at their preferred time. Another point would be to reduce the number of patient-reported measures or items, especially after discharge, when the motivation to respond diminishes. Adequately informing the staff and hierarchy could foster a positive attitude among the medical staff, a point agreed upon by all five study nurses as a facilitator. Implementation literature emphasizes the importance of informing and training staff, leadership engagement, as well as organizational support to promote staff acceptance and ensure the sustained integration into clinical workflows [23,48].

### Strengths and limitations

There are some potential limitations that should be considered when interpreting these findings. Results were embedded in a study context, limiting transferability to routine care. The study took place in GIM divisions of five Swiss university hospitals, which have varying infrastructures, workflows, local information technology systems, digital readiness, prior exposure to PROMs and PREMs and staff engagement. These settings have more resources and dedicated nurse support, limiting comparability and generalization to non-university hospitals. Furthermore, the limited number of study nurses (n=5) restricted the diversity of perspectives captured. Another limitation is that the AIM, IAM and FIM were administered to patients globally, not for each instrument individually, preventing patient-specific feedback. Finally, participation rates may not reflect willingness to complete PROMs and PREM in general.

This study has several notable strengths. First, the inclusion of five university hospitals enhances the external validity of the findings by capturing a high level of institutional diversity. Second, the dual perspective of patients and study nurses, combined with the use of both quantitative and qualitative methods, provides a comprehensive, multi-angle evaluation of PROM and PREM implementation. Third, the application of a validated set of implementation outcome measures [34] to assess acceptability, appropriateness, and feasibility strengthens the methodological rigor of the study. Finally, given the limited existing research on PROMs and PREMs in GIM populations living with multimorbidity, this study makes an important and novel contribution to literature.

## Conclusion

PROM and PREM use appear to be acceptable, appropriate, and feasible in a population of GIM inpatients living with multimorbidity, from the perspective of patients and study nurses. Most of the identified barriers were not population-specific, supporting the feasibility in GIM patients. However, multi-level support is required. The findings of this study can inform future research and implementation efforts, highlighting the need for further investigation in this under-documented setting and for linking current evidence to practical implementation processes.

## Supporting information

Supplementary materials with description

## Data Availability

Meta data are available from the corresponding author upon reasonable request.

## Acknowledgements

The authors would like to thank all the patients and study nurses for their participation, and Fiona Kraege for the questionnaire translation.

## CREDIT author statement

Conceptualization: M.M and C.S; investigation, data curation, formal analysis, visualization, draft writing: C.S; supervision, project administration, writing: M.M; review and editing, methodology: C.E.A, C.S, L.C, M.E; review and editing; J.R, M.E, J.S, S.B, F.V, L.C, V.K.

## Declaration of Generative AI and AI-assisted technologies in the writing process

No generative AI technologies were used in the writing of this manuscript, except for minor English language editing and formatting. In addition, the automated translation tool DeepL (DeepL SE, Cologne, Germany), which uses AI-based neural machine translation models specialized for language translation, was used to translate questionnaire materials. The authors take full responsibility for the content of the manuscript.

