## Supplementary materials with description for "Acceptability, Appropriateness and Feasibility of PROM and PREM Use in Swiss General Internal Medicine Divisions: A descriptive, sequential mixed method study"

**SUPPLEMENTARY MATERIAL**

**Supplemental Material 1.**

English version of the customized PREM used in the German and French surveys, translated using the automated translation tool DeepL (DeepL SE, Cologne, Germany).

English version of the customized PREM used in German and French, translated using *Deepl*.

**Questionnaire for the "TRADUCE" study**

**1.** In your opinion, did you receive any treatments (medication, physical therapy, urinary catheter, etc.) and/or tests (radiological imaging, blood tests, etc.) that you did not consider necessary?

 Yes  No

If yes: Can you indicate, in the categories below, which treatment or investigation you considered/felt was unnecessary? (multiple answers possible):

 Heart examination (cardiac ultrasound, coronary angiography*, ECG**)

*Coronary angiography is an examination that allows the arteries of the heart to be visualized using a contrast agent injected into the vessels.

** An ECG is an electrocardiogram where electrodes are placed on your chest.

 Radiological imaging (MRI, CT scan, scintigraphy, PET scan)

 Biopsy

 Procedure (surgical, colonoscopy*, gastroscopy**, bronchoscopy***, other procedure)

*Insertion of a tube into the rectum to view the inside of the colon using a camera on a screen.

**Insertion of a tube into the stomach to view the inside using a camera on a screen.

***Insertion of a tube into the lungs to view the inside using a camera on a screen.

 Other procedures (blood transfusion, insertion of a venous catheter, insertion of a urinary catheter, insertion of a gastric tube)

 Laboratory tests (blood sampling, urine collection, fingerstick blood glucose testing)

 Drug treatment (tablets)

 Drug treatment (intravenous infusion)

 Oncological treatment (chemotherapy, immunotherapy, radiotherapy, other cancer treatment)

 Physiotherapy treatment

 Other: ………………………………………….

**2.** Were you able to ask why certain tests were performed or certain treatments were prescribed?

 Yes  No

If yes: Did you feel comfortable asking the question?

 Yes  No

If not: why? (multiple answers possible):

 Lack of time (on your part)

 Lack of time on the part of medical staff

 You did not consider it important

 You did not have enough information

 You did not have enough energy for it

 You did not have the necessary knowledge/understanding

 Lack of self-confidence or courage / You felt intimidated

**3.** Would you like to share an experience that you were unable to describe in the previous questions about your care? Describe in 2-3 sentences what you would like to share with us.

(In order to take your experience into consideration and analyze it, please do NOT include any proper names (e.g.

my nurse, Solène Dupont) or any means of identifying staff (my caregiver, a young man with a tattoo on his cheek and red hair).

Free text experience: ……………………………………………………………………………

**4.** Would you like to make any suggestions for improvement regarding your care? Describe in 2-3 sentences what you would like to share with us.

(In order to take your experience into account and analyze it, please do NOT include any proper names (e.g., my nurse, Solène Dupont) or any means of identifying staff (my caregiver, a young man with a tattoo on his cheek and red hair).

Free text improvement : …………………………………………………………………………

5. Questionnaire evaluation :

Do you have any suggestions for improving this questionnaire? ………………………………

How long did it take you? ………………………………………………………………………

Were there any words that were difficult to understand? If so, which ones?

…………………………………………………………………………………………………..

**Supplemental Material 2.** Study procedures of the TRADUCE study

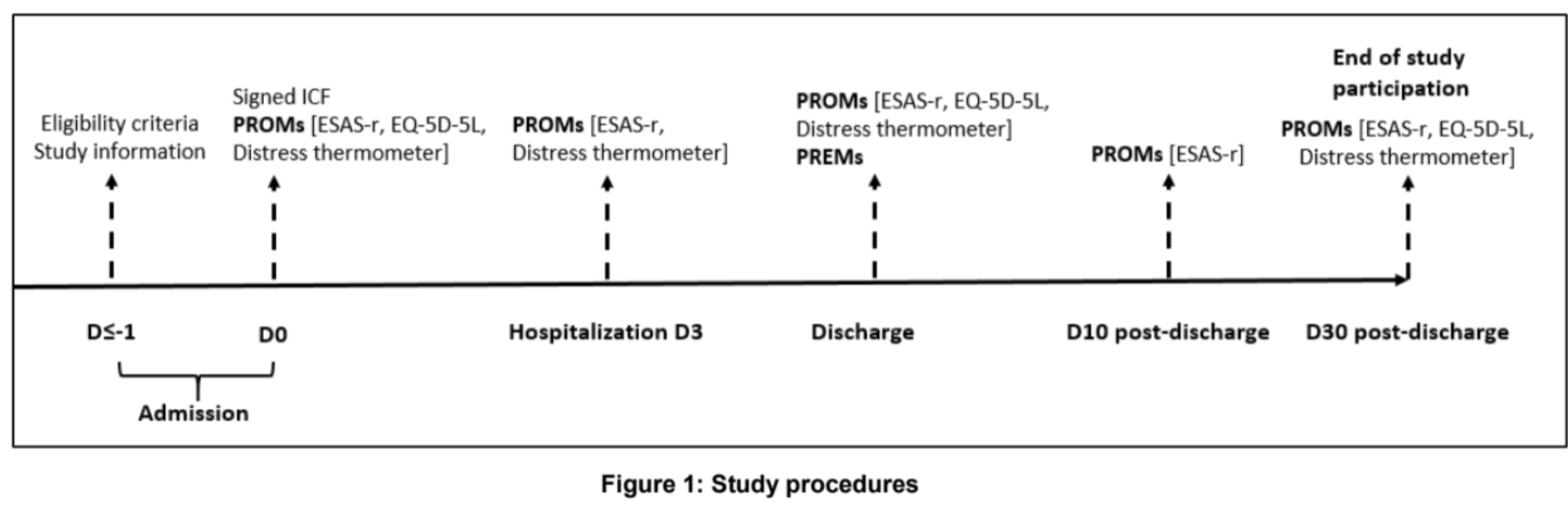

**Supplemental Material 3.** PROMs and PREM local source system used by the five hospitals of the TRADUCE study and special features

| **Hospital** | **Local source system** | **Special features** |
| --- | --- | --- |
| **Basel** | HEARTBEAT |  |
| **Bern** | REDCap |  |
| **Geneva** | Paper | Computerisation of scales in CONCERTO impossible for the study |
| **Lausanne** | ENGAGE |  |
| **Zurich** | REDCap |  |

**HEARTBEAT** (<https://heartbeat-med.com>): digital secure platform dedicated to PROMs and the collection of real data in clinical practice.

**REDCap** (<https://projectredcap.org>): secure system of research data collection used in clinical trials, observational studies and quality projects.

**CONCERTO** (Hôpitaux universitaires de Genève): secure institutional application used for collecting clinical and research data for hospital projects.

**ENGAGE** (<https://engagehelp.longenesis.com>): digital platform for participants and patients engaged in clinical trials and research projects.

**Supplemental Material 4.** Acceptability, appropriateness and feasibility of intervention measures: opinion survey in English for patients

**Opinion survey about the TRADUCE study**

Dear Patient,

You recently took part in a study called TRADUCE. During the trial you completed several questionnaires.

Three of the questionnaires were about your symptoms, possible feelings of distress and quality of life and are called **Patient Reported Outcome Measures** (**PROMs**). One questionnaire was about your experience of hospital care and is called the **Patient Reported Experience Measures** (**PREMs**).

We need to know how this experience was for you. That is why we are contacting you today.

Would you be willing to take part in a short 12-question survey to find out whether you think the PROMs and PREMs questionnaires are acceptable, appropriate and easy to use?

**YES**	**NO**

Thank you in advance for taking part.

**Acceptability of PROMs and PREMs of the TRADUCE study**

|  | Completely disagree | Disagree | Neither agree nor disagree | Agree | Completely agree |
| --- | --- | --- | --- | --- | --- |
| 1. PROMs and PREMs meet my approval. | ➀ | ➁ | ➂ | ➃ | ➄ |
| 2. PROMs and PREMs are appealing to me. | ➀ | ➁ | ➂ | ➃ | ➄ |
| 3. I like the PROMs and PREMs. | ➀ | ➁ | ➂ | ➃ | ➄ |
| 4. I welcome the PROMs and PREMs. | ➀ | ➁ | ➂ | ➃ | ➄ |

**Appropriateness of PROMs and PREMs of the TRADUCE study**

|  | Completely disagree | Disagree | Neither agree nor disagree | Agree | Completely agree |
| --- | --- | --- | --- | --- | --- |
| 1. PROMs and PREMs seem fitting. | ➀ | ➁ | ➂ | ➃ | ➄ |
| 2. PROMs and PREMs seem suitable. | ➀ | ➁ | ➂ | ➃ | ➄ |
| 3. PROMs and PREMs seem applicable. | ➀ | ➁ | ➂ | ➃ | ➄ |
| 4. PROMs and PREMs seem like a good match together. | ➀ | ➁ | ➂ | ➃ | ➄ |

**Feasibility of PROMs and PREMs of the TRADUCE study**

|  | Completely disagree | Disagree | Neither agree nor disagree | Agree | Completely agree |
| --- | --- | --- | --- | --- | --- |
| 1. PROMs and PREMs seem implementable. | ➀ | ➁ | ➂ | ➃ | ➄ |
| 2. PROMs and PREMs seem possible. | ➀ | ➁ | ➂ | ➃ | ➄ |
| 3. PROMs and PREMs seem doable. | ➀ | ➁ | ➂ | ➃ | ➄ |
| 4. PROMs and PREMs seem easy to use. | ➀ | ➁ | ➂ | ➃ | ➄ |

**Supplemental Material 5.** Acceptability, appropriateness and feasibility of intervention measures: opinion survey in French for patients

**Enquête d’opinion au sujet de l’étude TRADUCE**

Cher-e Patient-e,

Vous avez récemment participé à une étude intitulée TRADUCE. Au cours de l’étude, vous avez complété plusieurs questionnaires.

Trois des questionnaires concernaient vos symptômes, votre possible sentiment de détresse et votre qualité de vie et sont nommés **Patient Reported Outcome Measures** (ou mesures de résultats rapportés par le patient) (**PROMs**).

Un questionnaire portait sur votre expérience des soins hospitaliers et se nomme le **Patient Reported Experience Measures** (ou mesures de l’expérience de soins, rapportée par les patients) (**PREMs**).

Nous avons besoin de savoir comment cela s’est passé pour vous. C’est pourquoi nous vous contactons aujourd’hui.

Seriez-vous d’accord de participer à une courte enquête de 12 questions, afin de découvrir si vous pensez que les questionnaires PROMs et PREMS sont acceptables, appropriés et faciles à utiliser?

**OUI**	**NON**

Merci d’avance pour votre participation.

**Acceptabilité des questionnaires PROMs et PREMs de l’étude TRADUCE**

|  | Pas du tout d’accord | Pas d’accord | Ni d’accord ni pas d’accord | D’accord | Complètement d’accord |
| --- | --- | --- | --- | --- | --- |
| 1. J’approuve les PROMs et PREMs. | ➀ | ➁ | ➂ | ➃ | ➄ |
| 2. Les PROMs et PREMs suscitent mon intérêt. | ➀ | ➁ | ➂ | ➃ | ➄ |
| 3. Les PROMs et PREMs me plaisent. | ➀ | ➁ | ➂ | ➃ | ➄ |
| 4. J’accueille positivement les PROMs et PREMs. | ➀ | ➁ | ➂ | ➃ | ➄ |

**Pertinence des questionnaires PROMs et PREMs de l’étude TRADUCE**

|  | Pas du tout d’accord | Pas d’accord | Ni d’accord ni pas d’accord | D’accord | Complètement d’accord |
| --- | --- | --- | --- | --- | --- |
| 1. Les PROMs et PREMs semblent adaptés. | ➀ | ➁ | ➂ | ➃ | ➄ |
| 2. Les PROMs et PREMs semblent adéquats. | ➀ | ➁ | ➂ | ➃ | ➄ |
| 3. Les PROMs et PREMs semblent utilisables. | ➀ | ➁ | ➂ | ➃ | ➄ |
| 4. Les PROMs et PREMs semblent bien assortis. | ➀ | ➁ | ➂ | ➃ | ➄ |

**Faisabilité des questionnaires PROMs et PREMs de l’étude TRADUCE**

|  | Pas du tout d’accord | Pas d’accord | Ni d’accord ni pas d’accord | D’accord | Complètement d’accord |
| --- | --- | --- | --- | --- | --- |
| 1. Les PROMs et PREMs semblent implémentables. | ➀ | ➁ | ➂ | ➃ | ➄ |
| 2. Les PROMs et PREMs semblent plausibles. | ➀ | ➁ | ➂ | ➃ | ➄ |
| 3. Les PROMs et PREMs semblent faisables. | ➀ | ➁ | ➂ | ➃ | ➄ |
| 4. Les PROMs et PREMs semblent faciles à utiliser. | ➀ | ➁ | ➂ | ➃ | ➄ |

**Supplemental Material 6.** Online survey for study nurses

**A.** Acceptability, Appropriateness and Feasibility of Intervention Measures

| **Measure** | **Quizz** | **Questions** |
| --- | --- | --- |
| ESAS-r | Acceptability | ESAS-r meets my approval. |
|  |  | ESAS-r is appealing to me. |
|  |  | I like the ESAS-r. |
|  |  | I welcome the ESAS-r. |
|  | Appropriateness | ESAS-r seems fitting. |
|  |  | ESAS-r seems suitable. |
|  |  | ESAS-r seems applicable. |
|  |  | ESAS-r seems like a good match. |
|  | Feasibility | ESAS-r seems implementable. |
|  |  | ESAS-r seems possible. |
|  |  | ESAS-r seems doable. |
|  |  | ESAS-r seems easy to use. |
| EQ-5D-5L index | Acceptability | EQ-5D-5L index meets my approval. |
|  |  | EQ-5D-5L index is appealing to me. |
|  |  | I like the EQ-5D-5L index. |
|  |  | I welcome the EQ-5D-5L index. |
|  | Appropriateness | EQ-5D-5L index seems fitting. |
|  |  | EQ-5D-5L index seems suitable. |
|  |  | EQ-5D-5L index seems applicable. |
|  |  | EQ-5D-5L index seems like a good match. |
|  | Feasibility | EQ-5D-5L index seems implementable. |
|  |  | EQ-5D-5L index seems possible. |
|  |  | EQ-5D-5L index seems doable. |
|  |  | EQ-5D-5L index seems easy to use. |
| Distress thermometer | Acceptability | Distress thermometer meets my approval. |
|  |  | Distress thermometer is appealing to me. |
|  |  | I like the Distress thermometer. |
|  |  | I welcome the Distress thermometer. |
|  | Appropriateness | Distress thermometer seems fitting. |
|  |  | Distress thermometer seems suitable. |
|  |  | Distress thermometer seems applicable. |
|  |  | Distress thermometer seems like a good match. |
|  | Feasibility | Distress thermometer seems implementable. |
|  |  | Distress thermometer seems possible. |
|  |  | Distress thermometer seems doable. |
|  |  | Distress thermometer seems easy to use. |
| PREM from the TRADUCE study, i.e. on quality of care and excess care | Acceptability | TRADUCE PREMs meet my approval. |
|  |  | TRADUCE PREMs is appealing to me. |
|  |  | I like the TRADUCE PREMs. |
|  |  | I welcome the TRADUCE PREMs. |
|  | Appropriateness | TRADUCE PREMs seem fitting. |
|  |  | TRADUCE PREMs seem suitable. |
|  |  | TRADUCE PREMs seem applicable. |
|  |  | TRADUCE PREMs seem like a good match. |
|  | Feasibility | TRADUCE PREMs seem implementable. |
|  |  | TRADUCE PREMs seem possible. |
|  |  | TRADUCE PREMs seem doable. |
|  |  | TRADUCE PREMs seem easy to use. |

**B.** Questions

| **Section** | **Quiz** | **Question** |
| --- | --- | --- |
| General We have created a specific questionnaire to collect your appreciation of the PROMS and PREM use in the TRADUCE study, we thank you to answer to the following questions: | Quiz 1 | How feasible do you think it is to implement PROMs and PREMs routinely in internal medicine wards? |
|  |  | A. Very feasible |
|  |  | B. Moderately feasible |
|  |  | C. Slightly feasible |
|  |  | D. Slightly difficult |
|  |  | E. Moderately difficult |
|  |  | F. Very difficult |
|  |  | G. Not feasible |
|  | Quiz 2A | Compared to oncology and surgical departments, how do you perceive the feasibility of implementing PROMs and PREMs in internal medicine? |
|  |  | NB: If you don’t have experience of PROMS/PREMs use in other departments, please do not answer. |
|  |  | A. Much easier |
|  |  | B. Easier |
|  |  | C. Slightly harder |
|  |  | D. Harder |
|  |  | E. Much harder |
|  | Quiz 2B | Name the reasons: |
|  |  | NB: If you don’t have experience of PROMS/PREMs use in other departments, please do not answer. |
|  |  | (Open-ended text box) |
|  | Quiz 3 | The institution where I work supports the routine implementation of PROMs and PREMs for hospitalized patients. |
|  |  | A. Strongly agree |
|  |  | B. Agree |
|  |  | C. Neither agree nor disagree |
|  |  | D. Disagree |
|  |  | E. Strongly disagree |
| Barriers and Facilitators | Quiz 4 | Identify what are the institutional (where you work) barriers to implementing PROMs and PREMs. (Select all that apply) |
|  |  | A. Lack of resources |
|  |  | B. Lack of training |
|  |  | C. Resistance to change |
|  |  | D. Technological limitations |
|  |  | E. Fragmented care system |
|  |  | F. Patients factors (lack of interest, disease, etc) |
|  |  | G. Other (please specify) |
|  | Quiz 5 | What are the main facilitators for the successful implementation of PROMs and PREMs in your institution? (Select all that apply) |
|  |  | A. Strong leadership support |
|  |  | B. Adequate formation |
|  |  | C. Increasing digitalisation |
|  |  | D. Positive attitude from medical staff |
|  |  | E. Need to consider these patients |
|  |  | F. Patients factors (motivation to help, disease, etc) |
|  |  | G. Other (please specify) |
|  | Quiz 6 | The medical staff, you work with, is engaged and motivated to use PROMs and PREMs. |
|  |  | A. Strongly agree |
|  |  | B. Agree |
|  |  | C. Slightly agree |
|  |  | D. Slightly disagree |
|  |  | E. Disagree |
|  |  | G. Strongly disagree |
|  | Quiz 7 | How useful do you believe PROMs and PREMs are for improving patient care in internal medicine? |
|  |  | A. Very useful |
|  |  | B. Moderately useful |
|  |  | C. Slightly useful |
|  |  | D. Slightly useless |
|  |  | E. Moderately useless |
|  |  | F. Not useful |
|  | Quiz 8 | The use of PROMs and PREMs positively impacts the workflow of medical staff in internal medicine. |
|  |  | A. Strongly agree |
|  |  | B. Agree |
|  |  | C. Slightly agree |
|  |  | D. Slightly disagree |
|  |  | E. Disagree |
|  |  | F. Strongly disagree |
|  | Quiz 9 | Implementing PROMs and PREMs brings significant benefits to the institution, where you work. |
|  |  | A. Strongly agree |
|  |  | B. Agree |
|  |  | C. Slightly agree |
|  |  | D. Slightly disagree |
|  |  | E. Disagree |
|  |  | F. Strongly disagree |
|  | Quiz 10 | The digitalization (using smartphone or tablet or computer) of PROMs and PREMs is advantageous for their implementation. |
|  |  | A. Strongly agree |
|  |  | B. Agree |
|  |  | C. Slightly agree |
|  |  | E. Slightly disagree |
|  |  | F. Disagree |
|  |  | G. Strongly disagree |
|  | Quiz 11A | How feasible is it to use digital tools for PROMs and PREMs in your hospital? |
|  |  | A. Very feasible |
|  |  | B. Moderately feasible |
|  |  | C. Slightly feasible |
|  |  | D. Slightly difficult |
|  |  | E. Moderately difficult |
|  |  | F. Very difficult |
|  | Quiz 11B | Explain your response: |
|  |  | (Open-ended text box) |
|  | Quiz 12A | 12. Maintaining follow-up with patients post-discharge from internal medicine for PROM and PREM completion is challenging. |
|  |  | A. Strongly agree |
|  |  | B. Agree |
|  |  | C. Slightly agree |
|  |  | D. Slightly disagree |
|  |  | E. Disagree |
|  |  | F. Strongly disagree |
|  | Quiz 12B | Explain your response: |
|  |  | (Open-ended text box) |
| Study Execution | Quiz 13 | How satisfied are you with the overall execution of the TRADUCE study in your hospital? |
|  |  | A. Very satisfied |
|  |  | B. Moderately satisfied |
|  |  | C. Slightly satisfied |
|  |  | D. Slightly dissatisfied |
|  |  | E. Moderately dissatisfied |
|  |  | F. Very dissatisfied |
|  | Quiz 14 | The support and resources provided for the study are adequate. |
|  |  | A. Strongly agree |
|  |  | B. Agree |
|  |  | C. Slightly agree |
|  |  | D. Slightly disagree |
|  |  | E. Disagree |
|  |  | F. Strongly disagree |
|  | Quiz 15A | Experience with Reporting Unusual Scores: Have you ever encountered a situation where a patient's questionnaire responses indicated a direct impact on their health or care, as described in the TRADUCE study protocol? Specifically, this refers to the following statement: |
|  |  | French: "Je serai informé.e ainsi que les personnes qui s’occupent de moi pendant mon séjour hospitalier, si certaines des réponses aux questionnaires pourraient avoir une incidence directe sur ma santé ou prise en charge." |
|  |  | German: "Sowohl ich als auch Fachpersonen (Pflegefachpersonen und Ärztinnen/Ärzte), welche mich während des Spitalaufenthalts betreuen, werden informiert, falls einige der Antworten der Fragebogen direkte Auswirkungen auf meine Gesundheit oder meine Behandlung haben könnten." |
|  |  | A. Yes |
|  |  | B. No |
|  | Quiz 15B | If yes, please describe the situation and the actions taken: |
|  |  | (Open-ended text box) |
|  | Quiz 16 | Please provide any suggestions you have for improving the implementation of PROMs and PREMs or the execution of the TRADUCE study. |
|  |  | (Open-ended text box) |
| Conclusion Thank you for completing this survey. |  | Please indicate your institution here : |
|  |  | A. Basel |
|  |  | B. Bern |
|  |  | C. Geneva |
|  |  | D. Lausanne |
|  |  | E. Zurich |

**Supplemental Material 7.** Guide for the semi-structured focus group with the 5 study nurses, held in English, timing – 1 hour

| **Component** | **Question** |
| --- | --- |
| **Introduction** | Welcome and introductions: the interviewer introduces herself and explains the purpose of the meeting. She invites participants to introduce themselves (by name, hospital). |
|  | Purpose of the focus group: to explore in depth the perceptions and experiences of research staff regarding PROMs and PREMs, for future use in GIM hospitalized patients. |
|  | Rules of conduct: the interviewer ensures confidentiality of the discussion, encourages everyone to participate and to be respectful, explains the duration and progress of the session. Recording of the session to facilitate the analysis, material will be destroyed immediately after use. |
| **Advantages and disadvantages** | What do you think are the main advantages of PROMs/PREMs for GIM patients? |
|  | And for GIM healthcare professionals? |
|  | Have you encountered challenges or drawbacks/issues with the use of PROMs/PREMs in the TRADUCE study?  *If answer = yes:* what kind of issues, challenges? |
|  | Do you have any suggestions to overcome these challenges /issues? |
| **Implications for clinical practice** | How did patients accept PROMs/PREMs during the TRADUCE trial? |
|  | Did you receive feedback from healthcare providers (nurse, doctors) about PROMs/PREMs use?  *If answer = yes:* feedback was positive? Negative?  *If answer = yes:* can you give an example of feedback received? |
|  | Do you think all measure(s) should be implemented in clinical practice in GIM? |
|  | *If answer = no:* which among the 4 measures, ESAS, Distress, EQ5DL, PREMs would you prioritize? And why? |
| **Barriers and facilitators** | In the survey, you mentioned several barriers regarding PROMs/PREMs implementation in GIM, such as lack of resources, patient factors, …  How could they be reduced? |
|  | In the survey, you mentioned several facilitators regarding PROMs/PREMs implementation in GIM: dedicated teaching on PROMS and PREMs use, improving digitalization, positive attitude from medical staff.  How do you think you can promote them? |
| **Closing** | Ask if anyone has any further comments or questions. |
|  | Acknowledgements: for their time and valuable input. |

**Supplemental Material 8.** Analysis grid for the semi-structured focus group with the study nurses, used for MAXQDA qualitative analysis

|  | **Responses** | **Quotes** |
| --- | --- | --- |
| **1. Advantages and disadvantages** |  |  |
| Perceived advantages |  |  |
| For GIM patients |  |  |
| Sub-themes : |  |  |
| Key examples : |  |  |
| For GIM healthcare professionals |  |  |
| Sub-themes : |  |  |
| Key examples : |  |  |
| Disadvantages and challenges |  |  |
| Challenges encountered |  |  |
| Sub-themes : |  |  |
| Key examples : |  |  |
| Suggestions to overcome challenges |  |  |
| Sub-themes : |  |  |
| Key examples : |  |  |
| **2. Implications for clinical practice** |  |  |
| Adoption and engagement |  |  |
| Patient acceptance of PROMs/ PREMs during the TRADUCE trial |  |  |
| Sub-themes : |  |  |
| Key examples : |  |  |
| Feedback from healthcare providers |  |  |
| Sub-themes : |  |  |
| Key examples : |  |  |
| Measures |  |  |
| Measures from the TRADUCE study to be implemented in clinical practice in GIM |  |  |
| Sub-themes : |  |  |
| Key examples : |  |  |
| Measure(s) to prioritize and why |  |  |
| Sub-themes : |  |  |
| Key examples : |  |  |
| **3. Barriers and facilitators** |  |  |
| Barriers |  |  |
| Reducing barriers |  |  |
| Sub-themes : |  |  |
| Key examples : |  |  |
| Facilitators |  |  |
| Promoting facilitators |  |  |
| Sub-themes : |  |  |
| Key examples : |  |  |

**Supplemental Material 9.** Participation Rates on Digital Use, by Hospital.

| **Hospital** | **1** | **2** | **3** | **4** | **5** | **Overall** |
| --- | --- | --- | --- | --- | --- | --- |
| **Participation rate, n/N (%)** | 291/388 (75) | 244/457 (53) | 250/387 (65) | 204/408 (50) | 63/133 (47) | 1’052/ 1'773 (59) |
| **Digital n/N (%)** | 291/291 (100) | 112/244 (46) | 0/250 (0) | 19/204 (9) | 48/63 (76) | 470/1’052 (45) |

**Supplemental Material 10.** Qualitative display of semi-structured focus group with the study nurses

| **CONSTRUCT** | **Category** | **Qualitative coded answers** | **Qualitative reflective quotes (reproduced verbatim, including spelling and grammar)** |
| --- | --- | --- | --- |
| ADVANTAGES AND DISADVANTAGES | Advantages for GIM patients | expression of their feelings ; acknowledge the evolution of their state ; feeling of usefulness ; global picture on how they feel and its impact on their quality of life ; PREMs are useful to evaluate if the quality in the hospital is approved ; they give a vote about the patient's personal experience. | (N-4): « The patients could express what they feel, and what is good is that when they remember. So, they remember how they go through the pain, the fatigue etc., and its evolution. » (N-3): « Some of them feel useful also, to be useful to something in addition of only being hospitalized on the bed. » (N-1): « I think it's also for their discretioning the PREMs that are also useful for the quality management to prove if the quality in the hospital is approved. » (N-5): « I find it difficult, the digital use for all the multimorbit patients. » (N-2): « I think sometimes the questions were very private questions. Especially in the moments of distress. » |
|  | Advantages for healthcare professionals | a support for communication; efficiency and time saving ; brings up unusual conversations ; helps the patient to reflect on the personal course of treatment ; gives an overview about the patient and their whole social situation. | (N-4): « So an example of patient I've put three or four times; she was afraid she has not enough to eat at home. And this kind of conversation, they are not very usual with the physician or a nurse. » (N-1): « Because if the patient answers the PROMs and they have theses scores like nine fatigue or when they are tired, then you can start the conversation. » |
|  | Challenges encountered | digital use for multimorbid patients ; old patients need support to answer the questions ; tiredness and angriness of the patient ; difficulty for the patient to translate symptoms by numbers ; difficulty for the patient to use the right order of the scale ; mix up of the questionnaires timelines ; decrease of motivation to fill in the questionnaires ; too much questionnaires for some patients ; difficulty with the discharge questionnaire, as they already receive a satisfaction questionnaire and are eager to leave ; whole context for the patient makes it difficult to respond to the questionnaires ; questionnaires are not optimal for the GIM multimorbid patient ; difference between patients and their subjective scale ; interpretation of the responses ; issue about the availability of the patient ; the patient doesn't know exactly why they have to answer this questionnaire. | (N-2): « Not for the younger ones who quickly can do it by tablets, but for the more elder ones with paper and the ones that need support through the questionnaires. » (N-3): « And if they go home, it's difficult then to motivate them, or they forget. » (N-3): « I think if with the digital questionnaire it's easier, because with the paper I encountered an issue about the availability of the patient. Because they are the nurse, they are the physiotherapeuts, the interns. And then they go to the toilet, then the shower, then I mean etc. » |
|  | Suggestions to overcome challenges | directly transform the responses to the questionnaires into an usable score ; having validated questionnaires for the multimorbid patients of GIM ; reducing the number of questionnaires ; digitalisation ; not to keep the satisfaction questionnaire. | (N-4): « It should be recorded in one place, maybe with a score or something else, that the health care could see. » (N-3) : « I think also, I mean, if the objective is to use them in routine, it has to, yeah because I'm scared that the patient will, it's too much for them if it's done each time, because the polymorbits they come quite often at the hospital. » |
| IMPLICATIONS FOR CLINICAL PRACTICE | Patient acceptance of PROMs/ PREMs during the TRADUCE trial | better acceptance when there is only one questionnaire ; some questions are too private. | (N-5): « Some questions are perceived as too intimate, like sexual health or the ability to get children or spiritual questions. Or others are not understood properly, like, substance use or some points in the spiritual questionnaire. And they are a little bit overwhelmed about the many options. » |
|  | Feedback from healthcare providers | not a lot of contact with the healthcare providers ; nurses very supportive and found it very helpful, useful, to include patients with their opinion and their experiences ; advanced nurses are aware that PROMs and PREMs are coming soon ; chief nurses are very interested. | (N-5): « After my presentation, especially nurses were very supportive and found it very helpful, useful, to include patients with their opinion and their experiences into quality measurements. » |
|  | Measures of the TRADUCE study | EQ-5D-RL should be implemented ; ESAS is quite good but possibly long and confusing ; distress thermometer should be implemented ; depends on the illness and diagnosis and preference of the patient. | (N-3): « The quality of life it's the impact so that's right. It's really how they live with their disease and how it impacts their health. »  (N-3): « I think to communicate, the ESAS is good » (N-5): « Some questions are perceived as too intimate, like sexual health or the ability to get children or spiritual questions. Or others are not understood properly, like, substance use or some points in the spiritual questionnaire. And they are a little bit overwhelmed about the many options. » |
| BARRIERS AND FACILITATORS | Reducing barriers | digitalisation to be more precise with themselves ; digitalisation for availability of the patient ; try to find a good slot with the digital version ; find a place where they can be available to respond ; motivate them ; train the staff on how to communicate to the patients ; automatic answer to the PROMs ; explain directly the usefulness of the questionnaires for the patient. | (N-3): « Like to tell the professionals that it's really important to have the point of view of the patients, without questioning them directly actively, because they express different things when they do it by themselves. » (N-2): « And then I would say maybe motivate them that this would be something that better can contribute to their health. » (N-4): « It should be recorded in one place, maybe with a score or something else, that the health care could see. » (N-3) : « I think also, I mean, if the objective is to use them in routine, it has to, yeah because I'm scared that the patient will, it's too much for them if it's done each time, because the polymorbits they come quite often at the hospital. » |
|  | Promoting facilitators | help of the hierarchy ; prepare the nurses ; communicate with the patient ; research with the nurses ; promote the advantages of PROMs and PREMs to the patient ; communicate with the healthcare providers ; promote the advantages of PROMs and PREMs to the healthcare providers ; train the healthcare providers. | (N-1): « I think they can be more reflected to themselves because they have to do it alone. » (N-3): « To train them and to explain both the patient and the healthcare providers and also the hierarchy. » |
